# Effectiveness of a Text Message Intervention Promoting Seat Belt Use Among Targeted Young Adults: A Randomized Clinical Trial

**DOI:** 10.1101/2022.02.03.22270375

**Authors:** Brian Suffoletto, Maria L. Pacella-LaBarabara, James Huber, Claire Bocage, M. Kit Delgado, Catherine McDonald

## Abstract

**Background:** Seat belt use is an effective way to reduce the morbidity and mortality of motor vehicle crashes. However, 1 in 10 adults do not always wear a seat belt, with the lowest use rates of seat belt use among young adults. Digital behavioral interventions could be useful to increase seat belt use, but remain under-developed. This randomized clinical trial tested the efficacy of a 6-week behavioral text message program promoting seat belt use among young adults.

**Methods:** A parallel, 2-group, single-blind, individually randomized clinical trial. Eligible individuals recruited from 4 Emergency Departments were aged 18 to 25 years who reported driving or being a passenger without always using a seat belt in the past 2-weeks. Two hundred eighteen participants completed the 2-week trial run-in phase were randomly assigned 1:1 to intervention: self-monitoring control. The intervention arm (n = 110) received *SaVE*, a 6-week automated interactive text message program including weekly seat belt use queries with feedback and goal support to promote consistent use of a seat belt. The control arm (n = 108) received identical weekly seat belt use queries but no additional feedback. The primary and secondary outcomes were the proportion of young adults reporting always wearing a seat belt at 6- and 12-weeks, collected via web-based self-assessments and analyzed under intent-to-treat models.

**Results:** The mean (SD) age was 21.5 (2.2) years, 137 (63%) were female, 110 (50%) were White, and 33 (15%) were Hispanic. The 6-week follow-up rate was 86.2% (n = 187), with no differential attrition. At 6-weeks, 39.1% (95% CI, 30.0%-48.9%) of intervention participants always wore a seat belt vs. 23.1% (95% CI, 15.6%-32.2%) of control participants (odds ratio, 2.13; 95% CI, 1.18-3.84; *P* = .01). The 12-week follow-up rate was 64.2% (n = 140), with no differential attrition. At 12 weeks, 42.7% (95% CI, 15.6%-32.2%) of intervention participants always wore a seatbelt vs. 32.3% (95% CI, 20.6%-44.0%) of control participants (odds ratio, 1.25; 95% CI, 0.67-2.31; *P*=.48).

**Conclusions:** Results of this randomized clinical trial demonstrated that an interactive text message intervention using goal support was more effective at promoting seat belt use among targeted young adults than self-monitoring at 6-weeks. These findings, if replicated, suggest a scalable approach to help improve seat belt use and reduce crash-related morbidity and mortality.

**Trial registration:** ClinicalTrials.gov Identifier: NCT03833713

## Introduction

In 2020, the US Department of Transportation National Highway Traffic Safety Administration estimated that 38,680 people died in motor vehicle crashes (MVCs), with more than half of all MVC fatalities involved unbelted drivers or passengers.^1^ Despite strong evidence that seat belt use can reduce risk of major injury^2^ and save lives^3^, recent estimates in 2020 suggest that an estimated 10% of vehicle occupants still do not wear seat belts.^4^ Young adult drivers and passengers aged 18-24 have the highest crash-related non-fatal injury rates of all adults and a relatively low rate of seat belt use compared to other age ranges.^5^

Evidence-based prevention programs to increase seat belt use among targeted groups of young adults are needed to reduce injuries and prevent deaths. Seat belt mandates are effective policy level interventions to reduce MVC fatalities^6^, but with individual variation in behavior and lack of universal primary seatbelt enforcement, individual-level interventions are needed. Brief in-person behavioral interventions targeting vehicle safety have shown to improve seat belt use among older adults^7^, but have not been implemented broadly to affect public health.

Mobile digital behavioral interventions offer advantages to in-person behavioral interventions in their portability and automation. Portability allows for delivery of behavioral support in the context of everyday life in temporal proximity to when behavioral decisions are being made, thus increasing salience.^8^ Automation allows for standardization of support materials and eliminates the need to hire and/or train staff to perform counseling. Mobile digital behavioral interventions may be especially useful for young adults given their high personal ownership and use of of smartphones.^9^

One especially useful mobile communication modality for delivering behavioral support is text messaging. Text messaging is easy to use, discreet, anonymous, and a preferred communication modality among young adults. There is solid evidence^10^, including work from our group^11^, that automated text message interventions incorporating goal support can reduce risk behaviors in young adults. It remains unknown whether an automated text message intervention can improve seat belt use in targeted groups of young adults.

This study examined the effectiveness of a text message program promoting seat belt use in a randomized clinical trial among a demographically diverse sample of young adults Emergency Department patients who screened positive for past 2-week inconsistent seat belt use. We chose to recruit from the Emergency Department as it may be, for many young adults, their only point of intersection with health care.^12^ The primary hypothesis was that participants in the intervention arm would be more likely to report seat belt use at 6-weeks post-randomization than participants in a self-monitoring control arm. We also examined whether effects are durable out to 12 weeks, whether select young adults characteristics moderated the effectiveness of the intervention, and whether there are differential effects by vehicle seat location.

## METHODS

### Trial Design

The study was a parallel 2-group individually randomized clinical trial among young adults with inconsistent seat belt use. This study compared the following two conditions: 1) an interactive text message intervention using goal support to promote consistent use of a seat belt, and 2) a text message–based self-monitoring control. Assessor-blinded self-reported outcomes were assessed at 6- and 12-weeks post-randomization. The study was pre-specified in the trial protocol and registered with clinicaltrials.gov (NCT03833713). Based on estimates from Sommers et al., 2013^7^, the study was powered to detect a treatment difference of 15%, where 25% of the intervention participants and 10% of the self-monitoring controls report always using a seat belt at 6-weeks follow-up. Therefore, we sought to recruit a randomized sample of 108 individuals per group (216 total), which would provide 80% power at 2-sided alpha=.05 under an intention-to-treat (ITT) convention. Results are reported according to the Consolidated Standards of Reporting Trials (CONSORT) reporting guidelines^13^. The study was funded by the National Highway Traffic Safety Administration (NHTSA) who had no role in the design of the trial or interventions and approved by the University of Pittsburgh and University of Pennsylvania Institutional Review Boards. The full trial protocol can be accessed in Supplemental Materials.

### Recruitment and Enrollment

Between December 2019 and August 2021, with the exception of April to August, 2020 (due to COVID restricitons), patients aged 18–25 years who presented to one of 4 EDs in Pennsylvania were eligible to participate if they (1) were medically stable, (2) spoke English, and (3) reported less than always seat belt use in the past 2 weeks. Patients were excluded if they reported (1) not owning a personal mobile phone with text messaging, (2) planned to change their phones in the next 3 months, or (3) had no plan to drive and/or ride in a vehicle in the next month. Recruitment occurred at times and on days when a research associate was available, providing a convenience sampling of screened patients. Immediately following informed consent, participants completed the baseline assessment. All screened patients who reported less than always seat belt use were offered a standard resources sheet for information on seat belt safety.

### Run-in and Randomization

Participants who completed the baseline assessment were instructed to text a study telephone number. Only phone numbers that matched the phone numbers provided during the enrollment process were eligible. Once this match was recognized, participants received several texts welcoming them to the study and describing the trial run-in phase. The run-in was used primarily to exclude individuals who did not interact with the text message program. Each Sunday at 4pm, participants received the text message: “*How often have you been a passenger or driver in a car the past week? 0=never; 1=a few times; 2=most days; 3=every day*”, and if they responded with a value 1,2, or 3, received: “*How often did you wear a seat belt? 0=never; 1=a few times; 2=most of the time; 3=every time*”. Only participants who respond to at least 50% of these queries over the 2-week run-in phase were randomized to intervention or control by a computer algorithm that automated random allocation in a 1:1 sequence. Random assignments were in blocks of 4 based on recruitment site and concealed from participants and research staff throughout the trial.

### The Safe Vehicle Engagement (SaVE) Intervention

*SaVE* is a fully automated, interactive text message program to promote consistent seat belt use. It targets key constructs of the Theory of Planned Behavior^14^, attempting to alter attitudes toward wearing a seatbelt, providing cues to action, and boosting self-efficacy. Consistent with self-regulation^15^ and goal setting^16^ theories, SaVE incorporates weekly check-ins of seat belt use with tailored feedback as well as goal commitment prompts for the coming week, feedback, and reminders. Communication within SaVE is grounded in best-practices for digital behavioral interventions^17^ including personalization of each weekly dialogue with the participants name and identification of message origin as “*The SaVE Team*”. Each message was drafted by a team of health behavioral scientists and written at an appropriate level of literacy (Flesch-Kincaid). SaVE software was run by the Office of Academic Computing at the University of Pittsburgh Medical Center.

On the day of randomization, participants allocated to SaVE received a series of welcome messages describing what to expect over the intervention period. For example: “*Welcome to the SaVE Study. For the next 6 weeks, we will help you set goals and provide you personalized feedback on your seat belt use*.” Participants were instructed that they can drop out of the *SaVE* program at any time by texting “*STOP*.” Each Sunday, following the 2 weekly queries, which were identical to those sent in the run-in, if a participant reported always seat belt use, they received a positive reinforcement message, such as: “*Way to go*!”. If they reported less than always seat belt use, they received a feedback message reframing the goal failure as an opportunity for a fresh start^18^, such as: “*Bummer. There is always next week*.” Independent of their past week report, participants received a goal commitment query: “*Would you be willing to commit to a goal to wear a seat belt every time this week*?” If a participant agreed, they received a positive reinforcement message, such as: “*Good choice. You got this!*” On weeks when an individual agreed to commit to a goal, on Wednesday at 4pm, they received a message reminder, such as: “*Friendly reminder about your goal to wear your seat belt this week*.”If they did not agree to commit to a goal, they received a fact either about the risks of not wearing a seatbelt or benefit to wearing a seat belt, such as: “*Quick fact: Wearing a seat belt can reduce your chance of getting injured by half*.” For all queries, missing responses were re-prompted once only and if missing, participants received: “*You must be busy. We will check in next week*.” At the completion of 6-weeks, SaVE participants received the message: “*This completes the SaVE text messaging. Thanks for participating*.”

### Self-Monitoring Control

To isolate the effects of SaVE ‘s active components from self-monitoring alone, participants allocated to the control arm received weekly vehicle and seat belt queries identical to those sent in the run-in (i.e. *How often have you been a passenger or driver in a car the past week?* and “*How often did you wear a seat belt?)* without receiving any feedback or goal support. At the completion of 6-weeks, self-monitoring control participants received the message: “*This completes the SaVE text messaging. Thanks for participating*.”

### Measures

The baseline survey was conducted in-person in the ED using a web-based questionnaire and was hosted on a secure server. Follow-up assessments at 6- and 12 weeks post-randomization were conducted via a web-based questionnaire that required the participant to enter a unique password and hosted on a secure server. Participants who did not complete the survey online within 2 weeks were contacted over telephone by research staff blind to treatment assignment. Participants were eligible to receive a total of $45 for participation in the study, including $15 for completing the baseline assessment battery, $15 for completing the 6-week follow-up assessments, and $15 for completed the 12-week follow-up assessments. Participants were not compensated for completing text messages queries.

At baseline, participants provided demographic information, past 2-week vehicle use and safety, past month alcohol and cannabis use, past year driving history, and seat belt-related cognitions. Demographics included age, sex, race, ethnicity, and student status. Items to measure recent seat belt use were adapted from NHTSA’s Motor Vehicle Occupant Safety Survey^19^. These included: “*In the past 2-weeks*…*how often have you driven a car*?”, *“…how often have you been a passenger in the front seat of a car*?”, and “*…how often have you been a passenger in the back seat of a car?”*. For each seat position, we asked: “*How often did you wear a seat belt*..*?” with* response options of “*never, a few times, most days, every day*”. To measure distracted driving, we adapted an item taken from NHTSA’s Survey on Distracted Driving Attitudes and Behaviors^20^: “*Over the past 2 weeks, how often did you type on your phone while you were driving and when the car was moving?*” with response options: “*never, a few times, most days, every day*”. To measure alcohol-impaired driving, we adapted an item taken from NHTSA’s National Survey on Drinking and Driving Attitudes and Behaviors^21^: “*What’s the most number of alcoholic drinks you’ve had prior to driving on any occasion in the past 2 weeks*?”.

Past year driving history was measured usng items adapted from NHTSA’s Naitonal Survey of Speeding and Unsafe Drving Attitudes and Behavior^22^: 1) “*How many times in the last 12 months have you received a traffic ticket? (Not including parking tickets)*”; and 2) “*As a driver of a car, have you been in a crash in the past 12 months*?”. Past month binge drinking was measured using NIAAA’s definition^23^: “Ho*w many days over the last month have you drank more than (3 drinks for women/ 4 drinks for men)*?” and past month cannabis use was measured using an item from the NM-ASSIST^24^: “*How many days over the last month have you used cannabis (marijuana, pot, hash, grass, etc)*?”.

Select cognitive constructs related to seat belt use were measured using Theory of Planned Behavior^14^ and the Health Belief Model^25^, including: 1) perceived norms (i.e. “*How often do your friends use their seat belt*?”, with response options: “*never; rarely; most of the time; always*”); 2) perceived danger of not wearing a seat belt (i.e. “*How dangerous is it to not wear a seat belt*?” with response options: “*none; somewhat; very; completely*”); 4) and perceived control (i.e. “*How much do you agree: I have complete control over whether I wear a seat belt*.”, response options: “*strongly disagree; disagree; somewhat agree; mostly agree; strongly agree*”). Finally, we asked: “*Check off all the reasons you have not used a seat belt*”, with response options: “*uncomfortable; forgot; don’t think they help; like freedom; other*.*”*

At 6-weeks, participants completed measures of past 2-week vehicle use and safety, seat belt-related cognitions, and usability. Past 2-week vehicle use and safety and seat belt cognitions measures were identical to those used at baseline. Our primary outcome at 6-weeks was “*always using a seat belt*”, defined as an individual reporting “*every day*” to the frequency of seat belt use for all vehicle positions (i.e. driver, front passenger; rear passenger) over the past 2 weeks, coded as 1 if “*every day*” was reported for all seat positions or coded as 0 if less than “*every day*” was reported for any seat position. For example, if an individual reported wearing a seat belt “*every day*” as a driver, but “*most days*” as a rear passenger, their outcome was coded as 0. We chose this as our primary outcome because the goal of the intervention was to support consistent seat belt use. Usability was measured with 2 questions: 1) “*Did you find the text message program helpful” with response options:* “*not at all; somewhat; very much*”; and 2) “*Would you recommend the program to others*?, with resposnse options: *“yes; no*”.

At 12-weeks, participants completed measures of past 2-week vehicle use and safety and seat belt cognitions. Past 2-week vehicle use and safety and seat belt cognitions measures were identical to those used at baseline and 6-weeks. A secondary outcome of “*always using a seat belt use*” at 12-weeks was defined using the same procedure as at 6-weeks. We also looked at “*always using a seat belt*” in the positions of driver, front passenger and back seat passenger separately at 6- and 12-weeks as well as relative increase or descreases in seat belt use over time.

### Statistical Analysis

Primary outcome analyses compared point prevalence “*always using a seat belt*” at 6 weeks post-randomization in study arms using the logit function in Stata, version 16.1 (StataCorp, Inc) using an intention-to-treat (ITT) paradigm. As pre-defined in the study protocol, if a participant was missing seat belt use outcome data at 6-weeks follow-up, we imputed it based on weekly text message reports, using the worst performance recorded in the weeks closest to weeks 5 and 6, providing the most conservative estimate of the missing outcome. Secondary outcome analyses compared point prevalence “*always using a seat belt*” at 12 weeks post-randomization in study arms using multiple imputation. Predictors in the models included sex, race, Hispanic ethnicity, college enrollment, and “*always using a seat belt*” at 6 weeks post-randomization. The final inference was combined from 20 sets of imputed data, as per recommendations.^26^

Estimated treatment effects are reported as odds ratios (ORs) with 95% confidence intervals (95% CI). Sensitivity of the findings to imputation for 6- and 12-week outcomes were assessed by conducting completed case analyses (CCA). To identify potential moderators of the treatment-outcome relationship, we examined interactions between treatment assignment and limited baseline variables. We conducted stratified outcome analyses by any variable with a significant univariate interaction term. To understand if there were differential effects by location in vehicle, we compared the proportion of seat belt use frequency across driver, front and rear passenger positions. All hypothesis tests were conducted at a 2-tailed α = .05 significance level.

## RESULTS

### Study Flow and Retention

**Figure 1** shows the flow of young adults through the study. Between December 3, 2019 and June 18, 2021, 1732 young adults were identified as potential participants based on age, 1352 were approached for screening and 702 were screened for eligibility. A total of 286 participants completed the baseline assessment and 218 participants were randomized (110 to the intervention arm and 108 to the control arm). At 6-weeks, the follow-up completion rate was 86.2% (n = 187 of 218), with no differential attrition between arms (25.5% intervention vs. 29.6% control; p=0.49). At 12-weeks, the follow-up completion rate was 64.2% (n = 140 of 218), with no differential attrition between arms (39.8% intervention vs. 31.8% control; p=0.22). A comparison of baseline characteristics between 6-week responders and non-responders (see **Supplemental Table 1)** showed that age, race, past year MVC, and past month binge drinking exceeded the threshold for a potential effect (p<0.2) and were included in a multivariate logistic regression model, which identified the following associations with missing outcomes: age (OR=1.38; 95% CI 1.12-1.71); past year MVC (OR=0.25; 95% CI 0.08-0.78); and past month binge drinking (OR=0.26; 95% CI 0.10-0.66), none of which resulted in differential attrition by study arm. Only past month binge drinking (OR=0.29; 95% CI 0.10-0.92) was significantly associated with 12-week outcome missingness.

**Figure 1:**
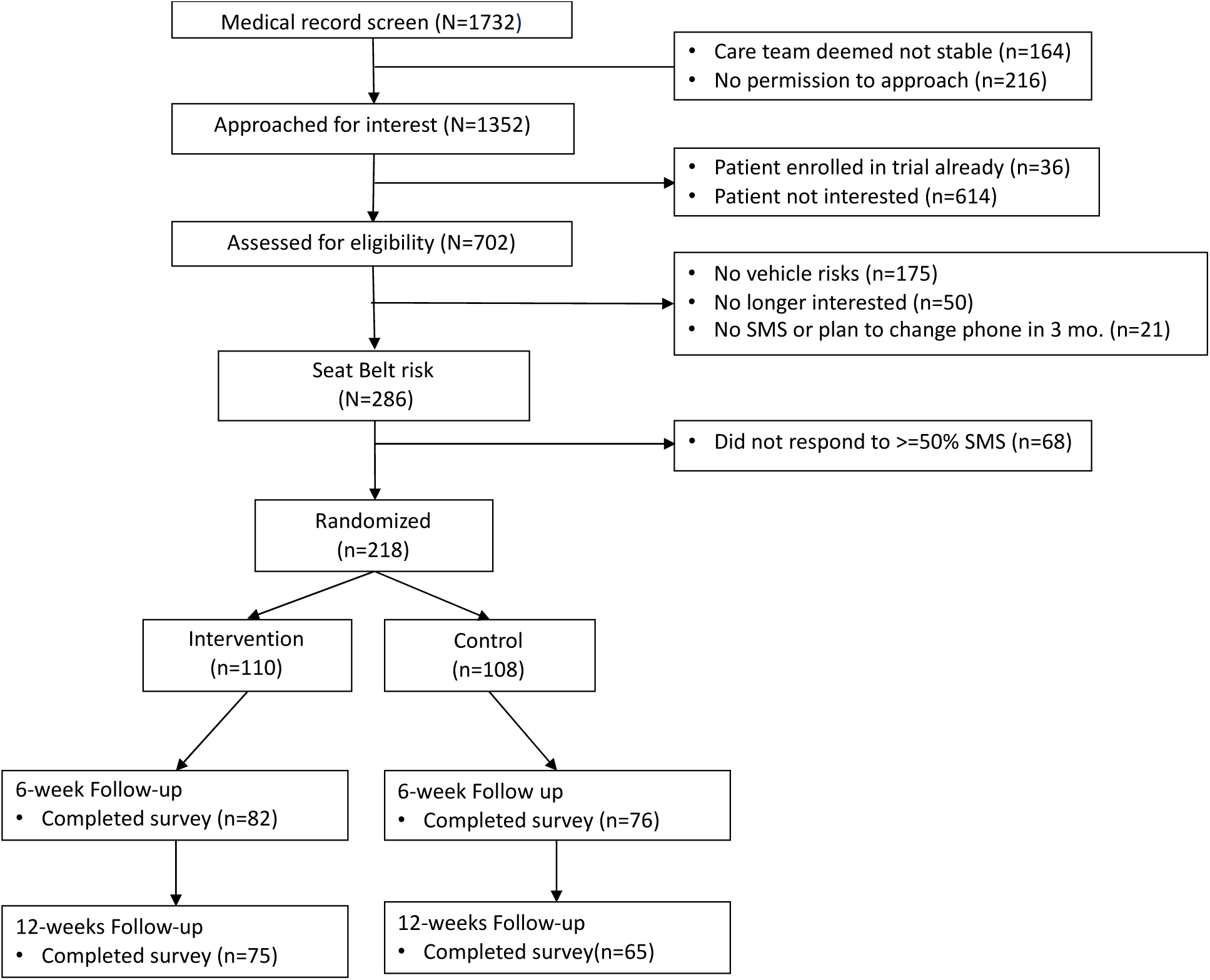
CONSORT Diagram.

### Participant characteristics

**Table 1** describes the self-reported baseline characteristics of enrolled participants. The mean (SD) of age of the 218 participants was 21.5 (2.1) years. The sample of participants was diverse in terms of sex (63.8% female), race (e.g. 33.0% Black), ethnicity (14.2% Hispanic) and education (37.6% currently enrolled in college). The majority of chief complaints for emergency care related to musculoskeletal pain (51.4%) or headache (26.2%). Only 3% of chief complaints were subsequent to MVCs. Participants reported high rates of vehicle activity across domains in the past 2 weeks, with 81.2% driving a car, 95.9% being a passenger in the front seat and 90.4% being a passenger in the back seat. The rate of always wearing a seat belt was lowest for back seat passengers (3.1%) and highest for drivers (46.9%). The majority of participants (65.6%) reported cellphone use while driving in the past 2 weeks and 8.7% reported driving a car after drinking alcohol. Over the past year, 15.6% reported receiving a traffic ticket as a driver and 14.5% reported being in an MVC. The most common reasons for not wearing a seat belt was forgetting (53.2%) and finding it uncomfortable (42.2%). Most participants (88.1%) perceived that their friends did not always wear a seat belt. Perceived danger of not wearing a seat belt and control over wearing a seat belt were both high. Over the past month, 35.8% reported binge drinking and 39.5% reported cannabis use. There were no significant (p value <0.05) between-arm differences in any baseline variables, indicating a balanced sample.

**Table 1.** Self-reported Baseline Characteristics of Enrolled Participants.

| **Characteristics** | **Total (N=218)** | **Intervention (n=110)** | **Control (n=108)** |
| --- | --- | --- | --- |
| Age, mean (SD), y | 21.5 (2.1) | 21.5 (2.2) | 21.6 (2.0) |
| Gender |  |  |  |
| Female | 139 (63.8) | 68 (61.8) | 71 (65.7) |
| Male | 79 (36.2) | 42 (38.2) | 37 (34.3) |
| Race |  |  |  |
| Black | 72 (33.0) | 37 (33.6) | 35 (32.4) |
| White | 111 (50.9) | 51 (46.4) | 60 (55.6) |
| Asian | 9 (4.1) | 7 (6.4) | 2 (1.9) |
| Mixed | 13 (6.0) | 5 (4.6) | 8 (7.4) |
| Other | 13 (6.0) | 10 (9.1) | 3 (2.8) |
| Hispanic | 31 (14.2) | 17 (15.5) | 14 (13.0) |
| Emergency Care Chief Complaint Category |  |  |  |
| MSK pain | 112 (51.4) | 62 (57.4) | 50 (45.4) |
| Headache | 57 (26.2) | 20 (18.5) | 37 (33.6) |
| Abd/Gyne/Uro | 11 (5.1) | 6 (5.6) | 5 (4.6) |
| Chest Pain/SOB/Syncope | 12 (5.5) | 4 (3.7) | 8 (7.3) |
| Other | 26 (11.8) | 16 (14.8) | 10 (9.1) |
| Current College enrollment | 82 (37.6) | 44 (40.0) | 38 (35.2) |
| **Past 2 week vehicle behaviors** |  |  |  |
| Drove car | 177 (81.2) | 88 (80.0) | 89 (82.4) |
| Always seatbelt use while driving | 83 (46.9) | 42 (47.7) | 41 (46.1) |
| Passenger in vehicle, front | 209 (95.9) | 105 (95.5) | 104 (96.3) |
| Always seatbelt use while front passenger | 75 (35.9) | 37 (35.2) | 38 (36.5) |
| Passenger in vehicle, back | 197 (90.4) | 99 (90.0) | 98 (90.7) |
| Always seatbelt use while back passenger | 6 (3.1) | 3 (3.0) | 3 (3.1) |
| Typing on phone when driving and car moving | 143 (65.6) | 71 (64.6) | 72 (66.7) |
| Drank alcohol prior to driving | 19 (8.7) | 7 (6.4) | 12 (11.1) |
| **Past year driving** |  |  |  |
| Traffic ticket as driver | 34 (15.6) | 13 (11.8) | 21 (19.4) |
| Motor vehicle accident as driver | 34 (14.5) | 10 (9.1) | 24 (22.2) |
| **Seat belt-related cognitions** |  |  |  |
| Reasons for not wearing a seat belt (not mutually exclusive) |  |  |  |
| Uncomfortable | 92 (42.2) | 50 (45.5) | 42 (38.9) |
| Forgot | 116 (53.2) | 63 (57.3) | 53 (49.1) |
| Don't think they help | 15 (6.9) | 7 (6.4) | 8 (7.4) |
| Like freedom | 28 (25.9) | 24 (21.8) | 28 (25.9) |
| Other | 26 (11.9) | 17 (15.5) | 9 (8.3) |
| Perceived friend use of seat belt, mean (SD) | 1.7 (0.8) | 1.7 (0.8) | 1.6 (0.8) |
| Perceived danger of not wearing a seat belt, mean (SD) | 2.5 (0.6) | 2.4 (0.7) | 2.6 (0.6) |
| Control over whether I wear a seat belt, mean (SD) | 3.4 (1.0) | 3.3 (1.1) | 3.5 (0.9) |
| **Substance use, past month** |  |  |  |
| Any binge drinking episode | 78 (35.8) | 36 (32.7) | 42 (38.9) |
| Any cannabis use | 86 (39.5) | 43 (39.1) | 43 (39.8) |

### Main Findings

**Table 2** shows the point prevalence of always using a seat belt at 6 and 12-weeks post-randomization in the study arms in addition to the rate difference and odds ratios using ITT and CCA. Rates of always seat belt use at 6-weeks under ITT were 39.1% (95% CI, 30.0%-48.9%) among intervention participants and 23.1% (95% CI, 15.6%-32.2%) among control participants (odds ratio, 2.13; 95% CI, 1.18-3.84; *P* = .01). Rates of always using a seat belt at 12-weeks under ITT were 42.7% (95% CI, 15.6%-32.2%) among intervention participants and 32.3% (95% CI, 20.6%-44.0%) among control participants (odds ratio, 1.25; 95% CI, 0.67-2.31; *P*=.48). CCA modeling confirmed similar estimates. The percentage of participants reporting different frequencies of seat belt use over time is shown in **Figure 2**.

**Table 2.** Seat belt Use Outcomes Under Intention-to-Treat and Complete-Case Analyses at 6- and 12-Weeks.

|  | Intervention arm | Control arm | Rate Difference | Odds ratio | P value |
| --- | --- | --- | --- | --- | --- |
| **6-weeks** | n=82 | n=76 |  |  |  |
| CCA | 41.5 (30.7 to 52.9) | 19.7 (11.5 to 52.9) | 21.8 (7.9 to 35.7) | 2.88 (1.41 to 5.89) | 0.03 |
|  | n=110 | n=108 |  |  |  |
| ITT | 39.1 (30.0 to 48.9) | 23.1 (15.6 to 32.2) | 16.0 (3.8 to 28.0) | 2.13 (1.18 to 3.84) | 0.01 |
| **12-weeks** | n=75 | n=65 |  |  |  |
| CCA | 42.7 (31.3 to 54.6) | 30.8 (19.9 to 43.4) | 11.9 (3.0 to 27.8) | 1.67 (0.83 to 3.36) | 0.15 |
|  | n=110 | n=108 |  |  |  |
| ITT | 42.7 (30.4 to 53.0) | 32.3 (20.6 to 44.0) | 10.4 (6.4 to 12.4) | 1.25 (0.67 to 2.31) | 0.48 |

**Figure 2.**
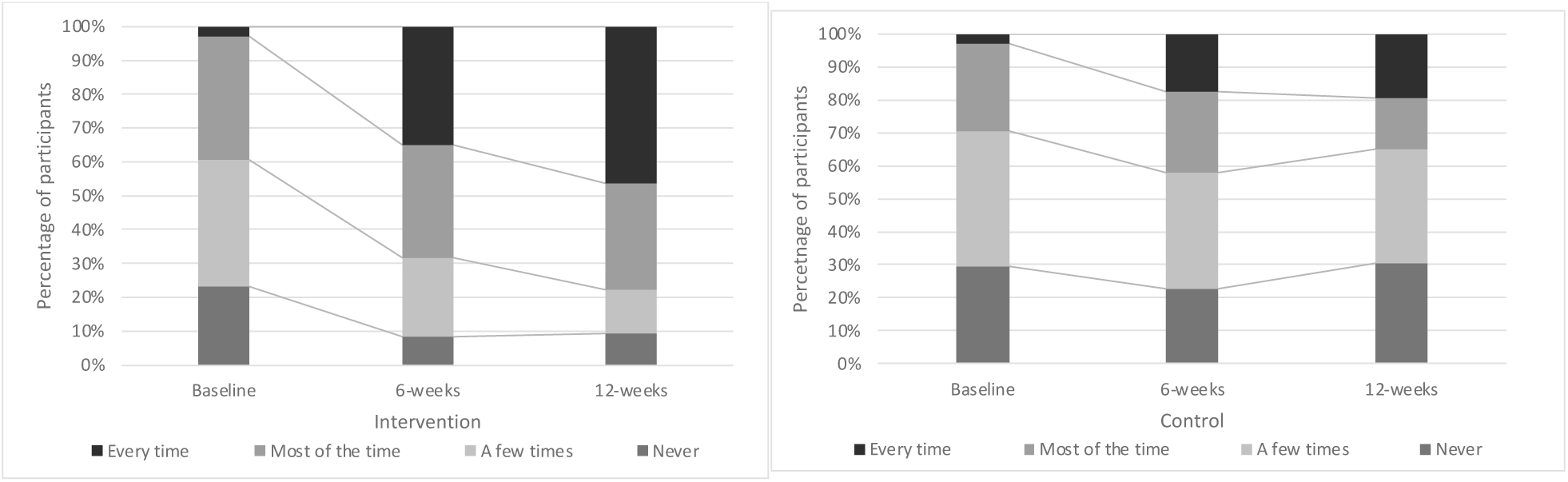
Change in Seat Belt Use over Time by Treatment Condition.

### Moderator Results

Analyses of baseline variables as potential moderators of treatment effects on point prevalence of always using a seat belt at 6-weeks post-randomization using ITT analysis identified perceived frequency of friend’s use of seat belts and past month cannabis use as having a significant interaction effects with treatment arm (see **Supplemental Table 2**). In stratified analyses, there were larger difference between Intervention and Controls when young adults perceived their friends using seat belts rarely or never compared with young adults who perceived their friends using seat belts most of the time or always (see **Table 3**). Also, difference between Intervention and Controls was larger among young adults who used cannabis compared to young adults who did not use cannabis.

**Table 3.** Moderators of Intervention Effects.

| **ITT 6-weeks: Always use seat belt** | Intervention arm | Control arm | Rate difference | Odds ratio | P value |
| --- | --- | --- | --- | --- | --- |
| Perceived friends use of seat belt |  |  |  |  |  |
| Mostly/Always | 38.5 (26.7-51.4) | 35.0 (23.1-48.4) | 3.5 (-13.4-20.4) | 1.16 (0.56-2.40) | 0.69 |
| Rarely/Never | 40.0 (25.7-55.7) | 8.3 (2.31-20.0) | 31.7 (21.1-54.2) | 7.33 (2.24-23.9) | 0.001 |
| **ITT 6-weeks: Always use seat belt** | Intervention arm | Control arm | Rate difference | Odds ratio | P value |
| Past month cannabis use |  |  |  |  |  |
| Yes | 46.5 (21.2-62.3) | 14.0 (5.3-27.9) | 32.5 (14.3-50.7) | 5.36 (1.88-15.3) | 0.002 |
| No | 34.3 (23.2-46.9) | 29.2 (18.6-41.8) | 5.1 (-10.8-20.8) | 1.27 (0.61-2.64) | 0.53 |

### Seat belt Use by Location in Vehicle

In examination of “always seat belt use” at 6- and 12-weeks post-randomization by location in vehicle identified using CCA, we found significant differences between Intervention and Control arms at 6-weeks and 12-weeks across driver, front and rear passenger positions. (see **Table 4**). At 6-weeks, the smallest difference between treatment arms was for back seat passengers and at 12-weeks the smallest difference between treatment arms was for front passengers.

**Table 4.**
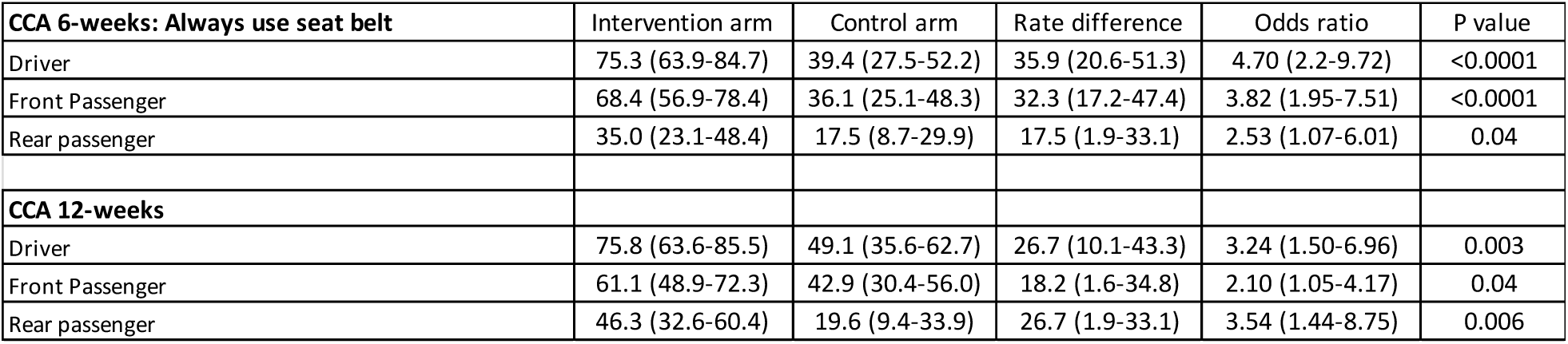
Intervention Effects by Vehicle Location.

| **CCA 6-weeks: Always use seat belt** | Intervention arm | Control arm | Rate difference | Odds ratio | P value |
| --- | --- | --- | --- | --- | --- |
| Driver | 75.3 (63.9-84.7) | 39.4 (27.5-52.2) | 35.9 (20.6-51.3) | 4.70 (2.2-9.72) | <0.0001 |
| Front Passenger | 68.4 (56.9-78.4) | 36.1 (25.1-48.3) | 32.3 (17.2-47.4) | 3.82 (1.95-7.51) | <0.0001 |
| Rear passenger | 35.0 (23.1-48.4) | 17.5 (8.7-29.9) | 17.5 (1.9-33.1) | 2.53 (1.07-6.01) | 0.04 |
| **CCA 12-weeks** |  |  |  |  |  |
| Driver | 75.8 (63.6-85.5) | 49.1 (35.6-62.7) | 26.7 (10.1-43.3) | 3.24 (1.50-6.96) | 0.003 |
| Front Passenger | 61.1 (48.9-72.3) | 42.9 (30.4-56.0) | 18.2 (1.6-34.8) | 2.10 (1.05-4.17) | 0.04 |
| Rear passenger | 46.3 (32.6-60.4) | 19.6 (9.4-33.9) | 26.7 (1.9-33.1) | 3.54 (1.44-8.75) | 0.006 |

### Usability

Two Intervention participants (1.8%) and no Control participants texted “STOP” during the 6-week intervention period. In the first 2 weeks of the intervention, 90.4% of participants completed the text message queries, which decreased to 77.5% by week 6. There were no significant differences in text message query response rates by Treatment arm. At 6-weeks follow-up, 49/81 (60.5%) of Intervention participants found the text message program very helpful compared to 27/73 (37.0%) of Control participants (p= 0.001). A total of 73/81 (90.1%) of Intervention participants would recommend the program to others compared to 63/73 (86.3%) of Control participants (p=0.46).

## DISCUSSION

This randomized trial provides the first experimental evidence that an automated and interactive text-message intervention focused on goal support can increase short-term seat belt use among a diverse sample of targeted young adults compared with a self-monitoring control. We found that, at 6-weeks, there was a 16% greater likelihood of participants randomized to SaVE to report always wearing their seat belt at the 6-week primary end-point compared with control participants. Based on this effect, 6 targeted young adults need to be exposed to the SaVE intervention to prevent one from being unrestrained in a vehicle. Estimates of the treatment benefit appear robust to assumptions about missing data, and by vehicle seat location.

Although there are no prior studies of seat belt digital interventions to compare to our study findings, in general, the effect size estimated in this study were comparable to other text message interventions for health promotion and harm reduction in young adults.^27^ Our findings can be contrasted with the only ED trial focused on seat belt safety, where an intervention including 1 face-to-face counseling intervention in the ED and a second telephone intervention 10 to 14 days after discharge resulted in increases in always using a seat belt from 53% at baseline to 59.5% at 12-weeks follow-up, a relative increase of 6.5%.^7^ This is much smaller than the relative increases we saw in our study in the intervention arm from 0% at baseline to 42.7% at 12-weeks follow-up but may be due to the higher baseline seat belt use rates in their study and the older population.

We found evidence of differential effects when examining each seat position separately where gretest differential effects at 6-weeks were for drivers and least effects for rear passengers, findings consistent with lower overall restraint use among rear passengers^28^ suggesting that there are false beliefs of safety, which could serve as a behavioral target for future interventions. We also found that the Control arm, which included weekly text queries about seat belt use, showed some improvement in seat belt use over time, with around 23% of participants reporting always wearing a seat belt at 6-weeks and 32.3% at 12-weeks. This finding fits with prior literature on the effects of self-monitoring on behavior change.^29(p)^

The superiority of the intervention was consistent across all demographic variables, yet seemed to be more effective among the subset of individuals who reported perceiving that their friends rarely or never use a seat belt and individuals who report past month cannabis use. The finding of differential effects by perceived peer norms fits with current understanding of the influence of peers on young adults driving behaviors^30^ and suggests that the text messages may either bolster individuals to act different from their peers or help correct distorted perceptions of peer behavior. The finding that the intervention may have stronger effects among individuals with cannabis use suggests that they may be more sensitive to intervention behavioral support. For example, the periodic nudges may help remind these individuals to wear a seat belt when they otherwise would not have remembered.

Strengths of this study include a diverse sample across a number of demographic characteristics (race, ethnicity, education) representative of the population of the US. Follow-up rates were higher than those in many text message studies conducted among young adults and there was no differential attrition across arms. Engagement with SaVE was high without financial incentives to do so. Given the low cost to send text messages and the automated nature of the intervention allowing deliver to almost every at-risk young adults in the US, it could have public health impact to reduce injuries related to unrestrained MVCs. For example, “prescriptions” for such a digital intervention could be bundled with ED discharge instructions or participation could be linked with driver’s insurance rates^31^. The feedback messages were based on decision rules that were developed prior to trial initiation, essentially eliminating the uncontrolled variability that exists in delivery of in-person interventions. The relative high fidelity to weekly text queries over 6 weeks and the high usability ratings suggest that the intervention may be acceptable to young adults outside a study setting.

Several potential limitations are worth noting. The outcome measures were based on self-reported data, which may be subject to recall or social desirability biases, and may have increased the apparent efficacy of the intervention. However, inclusion of a self-monitoring group, helped to guard against this possibility. This study did not include teens, in whom rates of seat belt use are poor. Future research should evaluate its effectiveness in this age group. Despite findings trends of differential effects, we were not powered to test significant effects at 12-weeks between Intervention and Control arms. Durable intervention effects could potentially be bolstered by running the Intervention longer than 6-weeks, consistent with behavioral literuate that it may take longer for behaviors to reach automoaticity.^32^ Finally, the trial was conducted during the unprecedented social disruption of the COVID-19 pandemic, which may have affected vehicle use and seat belt behaviors in unknown ways.

## Conclusions

This randomized clinical trial demonstrated the effectiveness of an automated, interactive text message intervention in promoting consistent seat belt use among young adults. These results establish a benchmark of effectiveness for other digital interventions aimed at improving vehicle safety and begin to fill an important gap in understanding how to help young people with inconsistent seat belt use who may not be exposed to vehicle safety support otherwise. A program like SaVE could fill a needed gap in supporting young adults to reduce the public health burden related to unrestrained MVCs.

## Data Availability

Data cannot be shared publicly because data is potentially identifiable. Data are available from the University of Pittsburgh Institutional Data Access / Ethics Committee for researchers who meet the criteria for access to confidential data.

---

**Supplemental Table 1.**
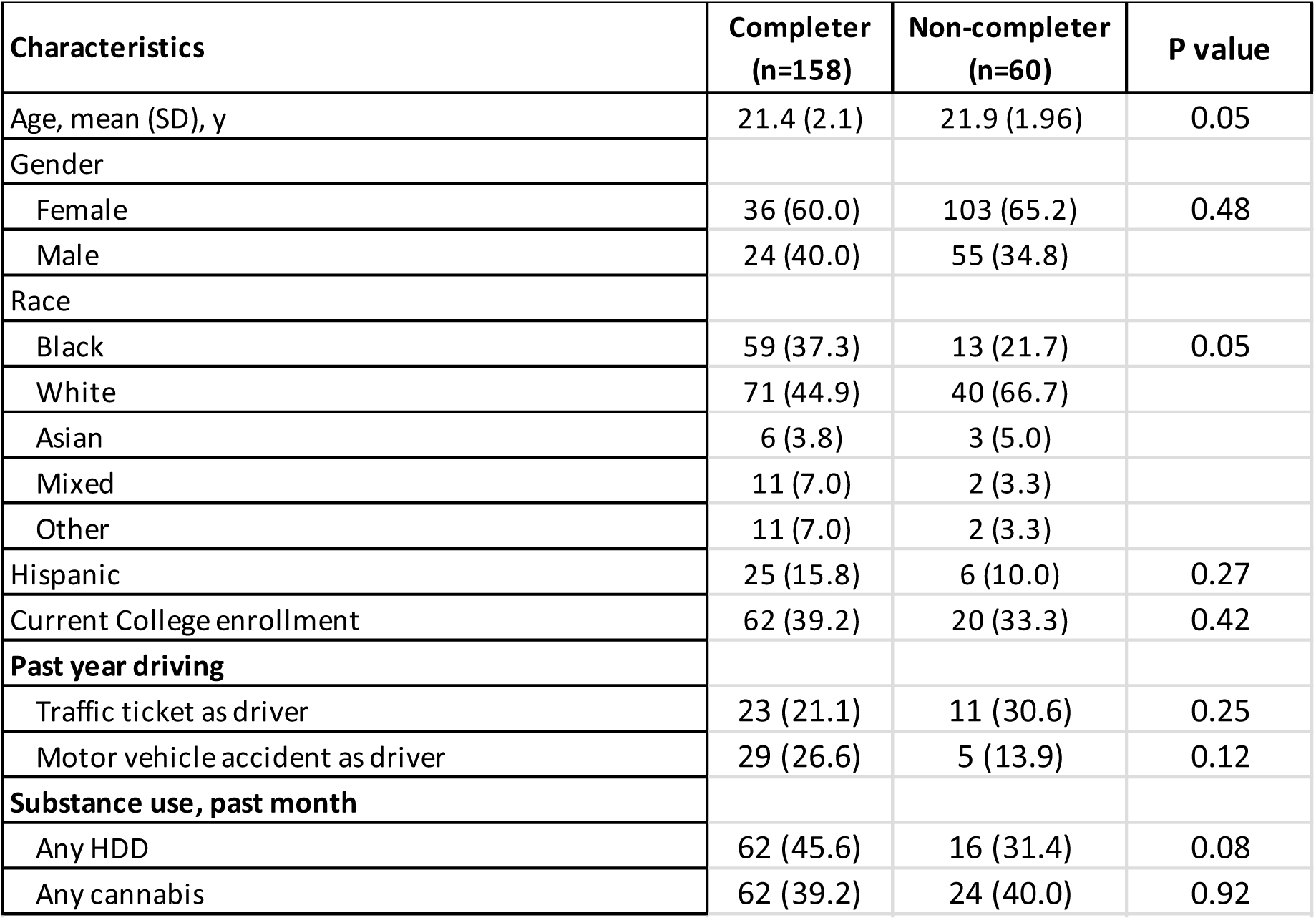
Comparison of Baseline Characteristics Between Non-Responders and Responders at 6-Weeks Follow-Up.

| Characteristics | Completer<br>(n=158) | Non-completer<br>(n=60) | P value |
| --- | --- | --- | --- |
| Age, mean (SD), y | 21.4 (2.1) | 21.9 (1.96) | 0.05 |
| Gender |  |  |  |
| Female | 36 (60.0) | 103 (65.2) | 0.48 |
| Male | 24 (40.0) | 55 (34.8) |  |
| Race |  |  |  |
| Black | 59 (37.3) | 13 (21.7) | 0.05 |
| White | 71 (44.9) | 40 (66.7) |  |
| Asian | 6 (3.8) | 3 (5.0) |  |
| Mixed | 11 (7.0) | 2 (3.3) |  |
| Other | 11 (7.0) | 2 (3.3) |  |
| Hispanic | 25 (15.8) | 6 (10.0) | 0.27 |
| Current College enrollment | 62 (39.2) | 20 (33.3) | 0.42 |
| **Past year driving** |  |  |  |
| Traffic ticket as driver | 23 (21.1) | 11 (30.6) | 0.25 |
| Motor vehicle accident as driver | 29 (26.6) | 5 (13.9) | 0.12 |
| **Substance use, past month** |  |  |  |
| Any HDD | 62 (45.6) | 16 (31.4) | 0.08 |
| Any cannabis | 62 (39.2) | 24 (40.0) | 0.92 |

**Supplemental Table 2.** Moderators of Intervention Effects.

| **Characteristics** | **Beta** | **Std. Error** | **P value** |
| --- | --- | --- | --- |
| Age, mean (SD), y | -0.10 | 0.15 | 0.51 |
| Gender (reference=Female) |  |  |  |
| Male | -0.17 | 0.62 | 0.79 |
| Race (reference=White) |  |  |  |
| Non-White | -1.10 | 0.61 | 0.08 |
| Hispanic ethnicity | -0.59 | 1.04 | 0.57 |
| Current College enrollment | -1.22 | 0.62 | 0.05 |
| **Past 2 week vehicle behaviors** |  |  |  |
| Typing on phone when driving and car moving | -0.61 | 0.65 | 0.34 |
| Drank alcohol prior to driving | 0.05 | 1.06 | 0.96 |
| **Past year driving** |  |  |  |
| Traffic ticket as driver | 0.38 | 0.86 | 0.66 |
| Motor vehicle accident as driver | 0.86 | 0.92 | 0.64 |
| **Seat belt-related cognitions** |  |  |  |
| Reasons for not wearing a seat belt (not mutually exclusive) |  |  |  |
| Uncomfortable | 0.85 | 0.63 | 0.18 |
| Forgot | 0.02 | 0.61 | 0.98 |
| Don't think they help | -0.61 | 0.85 | 0.89 |
| Like freedom | 1.33 | 0.76 | 0.08 |
| Other | -0.45 | 0.84 | 0.95 |
| Perceived friend use of seat belt, mean (SD) | -1.30 | 0.44 | <0.01 |
| Perceived danger of not wearing a seat belt, mean (SD) | -0.89 | 0.56 | 0.12 |
| Control over whether I wear a seat belt, mean (SD) | -0.40 | 0.38 | 0.29 |
| **Substance use, past month** |  |  |  |
| Any HDD | 0.48 | 0.65 | 0.46 |
| Any cannabis | 1.44 | 0.65 | 0.02 |

